# Field clinical performance of SARS-CoV-2 point-of-care diagnostic tests: a living systematic review of trials up to 17^th^ of August, 2021

**DOI:** 10.1101/2021.09.20.21263509

**Authors:** Gabriel Hawthorne, Adam Harvey

**Affiliations:** Cambridge University Hospitals NHS Foundation Trust, Cambridge, United Kingdom; Diagnostics for the Real World EU, Chesterford Research Park, Saffron Walden, United Kingdom

## Abstract

Point-of-care assays offer a decentralized and fast solution to the diagnosis of SARS-CoV-2, providing benefits for patients, healthcare workers and healthcare facilities. This technology has the potential to prevent outbreaks, enable fast adoption of potentially life-saving measures and improve hospital workflow. While reviews regarding the laboratory performance of those assays exist, a review focused on the real-life clinical performance and true point-of-care feasibility of those platforms is missing. Therefore, the objective of this study is to help clinicians, healthcare providers and organizations to understand the real-life performance of point-of-care assays, aiding in their implementation in decentralised, true point-of-care facilities, or inside hospitals. 1246 studies were screened in 3 databases and 87 studies were included, evaluating 27 antigen tests and 11 nucleic-acid amplification platforms deemed feasible for true point-of-care placement. We excluded studies that used processed samples, pre-selected populations, archived samples and laboratory-only evaluations and strongly favored prospective trial designs. We also investigated package inserts, instructions for use, comments on published studies and manufacturer’s websites in order to assess feasibility of point-of-care placement and additional information of relevance to the end-user. Apart from performance in the form of sensitivity and specificity, we present information on time to results, hands-on time, kit storage, machine operating conditions and regulatory status. To the best of our knowledge, this is the first review to systematically compare point-of-care test performance in real-life clinical practice. We found the performance of tests in clinical practice to be markedly different from the manufacturers reported performance and laboratory-only evaluations in the majority of scenarios. Our findings may help in the decision-making process related to SARS-CoV-2 test in real-life clinical settings.

**Rationale for the review:** A review focused on the real-life clinical performance and point-of-care feasibility of SARS-CoV-2 diagnostic platforms is missing, impairing the ability of individuals, healthcare providers and test providers to make informed decisions.

**Objective(s) or question(s) the review addresses:** The objective of this study is to help clinicians, healthcare providers and organizations to understand the real-life performance of point-of-care assays, aiding in their implementation in decentralised, true point-of-care facilities or in complex healthcare environments.

## Introduction

In December 2019, SARS-CoV-2 was first reported in Wuhan, China, and a pandemic was declared by the World Health Organization (WHO) in March, 2020. Reverse transcription-quantitative polymerase chain reaction (RT-qPCR) is the gold standard for diagnosis of SARS-CoV-2 infection. However, this technique has disadvantages including the requirement of centralised facilities with specific equipment, the requirement of highly trained staff and a long turnaround time between sample collection to results, approaching over 48 hours in some scenarios^1^. Driven by the need for diagnostic solutions during the pandemic, multiple new assays and platforms were developed, including multiple fast molecular and antigen tests. The FIND SARS-COV-2 DIAGNOSTIC PIPELINE^2^, which collates an overview of commercially available SARS-CoV-2 tests in real time, listed 1152 results for diagnostic solutions up to 29^th^ of October, 2021.

Point-of-care (POC) diagnostic platforms are defined as diagnostic systems that can deliver results near patients, without the need for centralized laboratories or diagnostic facilities^3^. These platforms tend to deliver fast results, often within 120 minutes, enabling rapid medical decisions and facilitating timely interventions. POC diagnostic assays are currently in use in health systems for different ends, from the bedside glucose test^4^ to the analysis of blood gases and electrolytes^5^. Besides being able to provide faster results, an important advantage of POC tests is to facilitate diagnosis in locations that previously could not have access to centralized laboratory diagnostic techniques. In the context of transmissible infectious diseases, some of these assays enable quick decisions regarding treatment and public health measures such as isolation of individuals and contact-tracing. Before the pandemic, POC tests were already in use for the diagnosis of conditions such as influenza-like illnesses in different settings, including accident and emergency departments in hospitals and outpatient clinics^6^. Other assays focus on the diagnosis of sexually transmitted diseases like HIV^7^ and Chlamydia trachomatis^8^. As a consequence of the SARS-CoV-2 pandemic and the need for more effective diagnostic solutions, not only centralized diagnostic solutions but also POC diagnostic assays had an unprecedent expansion, since time from sample collection to results is key to prevent further infections and to speed up patient triage in hospitals and healthcare settings overall.

However, POC tests can have limitations such as decreased sensitivity or specificity, increased costs, and lower throughput compared to centralised laboratory facilities and techniques such as real-time PCR. Some tests also require multiple manual steps in sample preparation or computers for their execution, which can make platforms too complex for true POC placement. Additionally, tests with poor performance can have multiple consequences. False negative results can cause inadequate placement of patient in hospitals (e.g, moving a infectious patient to a ‘green ward’), causing new outbreaks in an already diseased population, and also deem a community patient not infectious, thus increasing the risks of propagating infection to contacts. False positive results can inversely place patients in high-risk environments in hospitals (e.g, inside a ‘red ward’) and cause unnecessary isolation and economic impact in an outpatient setting. If a test is considered inaccurate and might require confirmation before results can be clinically acted on, this defeats the purpose of a rapid test.

The challenges of testing in real-life scenarios have also been explored. For instance, Micocci et al^9^ interviewed staff from English care homes and reported on the difficulties of COVID-19 testing in that setting. Isolation and testing procedures were found to be challenging, requiring reconfiguration of staffing and the environment. One of the conclusions of the study was that each POC test must be evaluated in the context the test is going to be conducted in, validating the need for in depth detail for the platform in question.

The regulatory aspect of novel tests in the context of a pandemic has been complex, and the extent of this process was likely unprecedented. Due to the urgent need for testing solutions, many abbreviated validation studies were accepted by the scientific community and by regulatory agencies with initial or partial evaluations, and few assays had their field performance assessed before being released to the market. Despite showing good accuracy in internal laboratory validations, multiple POC tests for SARS-CoV-2 diagnosis had a lower-than-expected performance once released for clinical use^10^. This topic was the subject of political and legal debate^11^ and resulted in some previously approved tests being later withdrawn from the market^12^. There are many examples of disparities between manufacturer claims based on laboratory-only evaluations and data from clinical trials. For instance, the platform ID Now (Abbott) claimed a sensitivity of 100% and a specificity of 100% on the product’s package insert^13^, but clinical evaluations showed a sensitivity below 75%^14^ or below 50% in some cases^15^. Another example is the study conducted by Jokela et al^16^, where the Mobidiag Novodiag platform had 93.4% (100/107) sensitivity in archived samples but only identified 60% (3/5) positive samples in a clinical setting.

POC diagnosis plays an important role in SARS-CoV-2 management. Faster diagnosis speeds up isolation measures and therefore the prevention of new outbreaks. In the same way, faster confirmation of SARS-CoV-2 absence helps avoiding unnecessary isolation for individuals and their contact groups, providing social and economic benefit. The workflow of patients inside a hospital can be greatly facilitated by using tests that are reliable and provide a fast result, aiding in the placement of patients inside “red” or “green” wards and preventing SARS-CoV-2 nosocomial spread while freeing up rooms and improving the capacity of emergency departments. For example, in the study conducted by Collier et al using the SAMBA platform^17^, mean length of stay on COVID-19 “holding” (or “amber”) wards was reduced by nearly 30h using the POC test. Additionally, timely interventions like the use of dexamethasone in patients requiring respiratory support^18^ or the use of interleukin-6 receptor antagonists in critically ill patients^19^ benefit from a fast diagnostic modality, especially considering the waiting-time for a centralised test result.

In this review, we address the POC tests for SARS-CoV-2 diagnosis, that are divided in two large categories. The first category is of the nucleic acid amplification tests (NAAT), also called molecular tests. These tests use some sort of nucleic acid amplification technique (polymerase chain reaction or a similar technique), and therefore require platforms for test conduction. The second category concerns antigen tests, which directly target SARS-CoV-2 surface antigens. These tests are usually attached to a lateral flow detection cassette and more often than not have no associated platform. We specifically focus on clinical trials clarifying the performance of those tests in real life settings. Our criteria was rigidly tailored to exclude samples that were pre-selected and testing conducted in laboratory conditions and/or with frozen samples. The objective of this study is to help clinicians, healthcare providers and organizations to understand field performance of POC assays, aiding in their safe implementation in decentralised, true POC facilities or in more complex healthcare environments.

Other reviews of POC assays targeting SARS-CoV-2 are available and use different inclusion criteria. Dinnes et al^20^ published a review that includes 64 studies for 16 antigen platforms and 5 molecular assays, collected up to September 2020. This study attempted to divide reports between symptomatic and asymptomatic individuals, and also included laboratory-only evaluations and retrospective studies, which diverges from our goal. Yoon et al^21^ conducted a study using the FDA Emergency Use Authorization (FDA-EUA) POC tests up to August, 2020, in which 26 studies were analyzed. Hayer et al^22^ reviewed antigen tests but only included assays not needing a separate reader. Other POC diagnostic technologies that have been in use in the management of COVID-19 patients, like the use of POC ultrasound for patient follow-up after diagnosis, do not fit the scope of this work.

## 1. Methodology

This systematic review was conducted following Preferred Reporting Items for Systematic Reviews and Meta-Analyses (PRISMA) guidelines.

After debate between the authors, a broad search phrase was defined with the intention to capture a large number of studies, given the novelty of the field. Filtering results by date using the abovementioned algorithm was not necessary, since the name SARS-CoV-2 was coined after the identification of the pathogen in 2019.

### 2.1 Search strategy

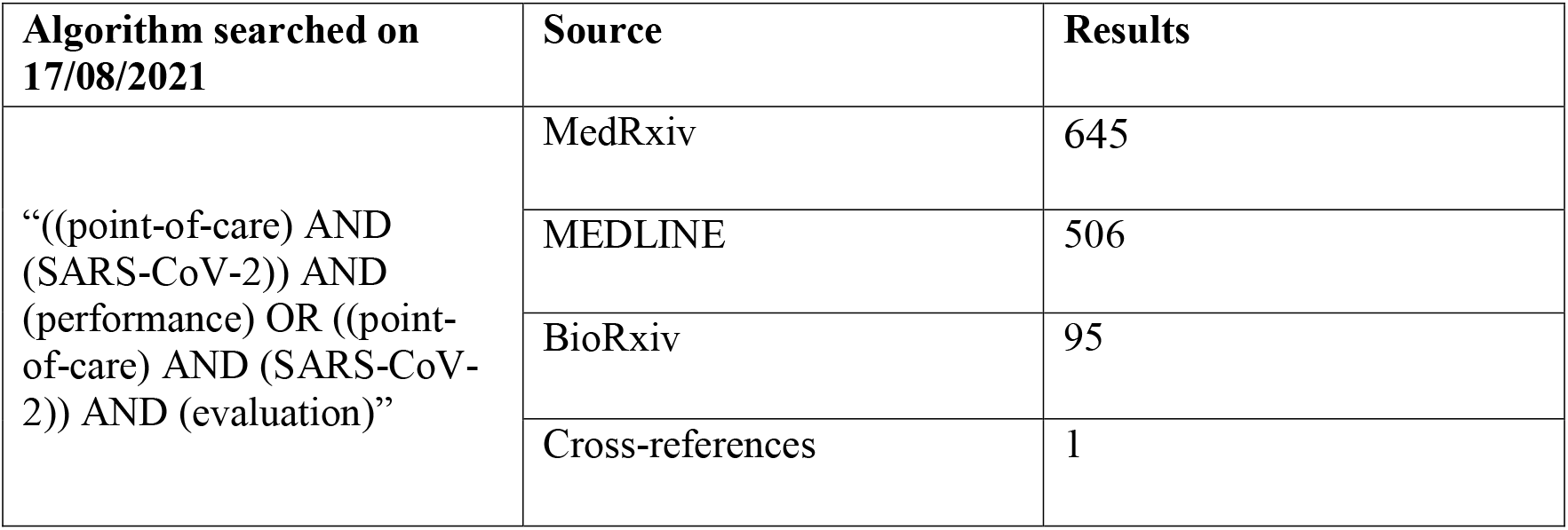

| Algorithm searched on<br>17/08/2021 | Source | Results |
| --- | --- | --- |
| “((point-of-care) AND (SARS-CoV-2)) AND (performance) OR ((point-of-care) AND (SARS-CoV-2)) AND (evaluation)” | MedRxiv | 645 |
|  | MEDLINE | 506 |
|  | BioRxiv | 95 |
|  | Cross-references | 1 |

### 2.2 Exclusion criteria

| Exclusion criteria |
| --- |
| -Study unrelated to POC testing; |
| -Platform not deemed feasible for the POC (e.g requires the use of centralised equipment); |
| -Test main focus other than diagnosis of acute infection (e.g antibody assays); |
| -Study not a clinical trial (e.g proof of concepts, validations, studies conducted with frozen samples, test conducted in laboratory conditions); |
| -Samples collected from patients processed before testing; |
| -Population pre-selected (e.g, only known positives were enrolled); |

### 2.3 Selection process

#### 2.31 Defining a POC assay

The definition of a POC diagnostic assay is not straightforward, since there are no rigid criteria for reference. The defining factor is being able to provide a diagnostic solution near the patient, thus removing the need for a centralized testing facility. While some platforms are totally mobile, being able to move to the patient’s location and clearly defined as POC, others require the use of an energy source, centralised computers, tablets or other accessories. Other assays require multiple sample preparation steps before test conduction (such as the need for RNA extraction, the use of a centrifuge, heating blocks or multiple pipetting steps), even though the final testing step can theoretically be conducted near the patient; this is the case of most LAMP platforms. In addition, requirements for isolation and measures for preventing infection spread further complicate this definition during the SARS-CoV-2 pandemic. We therefore recognize the definition of a POC test or platform remains subjective. Aligned with our objectives, in this review, we considered an outpatient community setting as our reference point. Therefore, assays that use reagents requiring processing in a central laboratory or facility were excluded from our analysis. The rationale for that is that these platforms could not be operational without such facilities in the vicinity and therefore could not be implemented in the community in a truly POC fashion. Based on the information available in publications and manufacturer’s manuals, assays that are estimated to be technically capable of implementation in lower complexity settings are included. These platforms will have their potential limitations/considerations (e.g, the requirement for cold chain storage or the need for a desktop computer) described in the comments of the results table.

#### 2.32 Defining target trials and comments regarding the criteria

Because of the disruption and urgency caused by the COVID-19 the pandemic, many studies have not followed rigid clinical trial designs, and a high level of heterogeneity between methods and designs is acknowledged. Issues such as need for self-isolation, patient discomfort upon repeated swabbing, multi-platform evaluations and insufficient number of positive samples in low-incidence scenarios affect the feasibility of the studies and need to be taken into consideration when conducting a review. For instance, the study of Tu et al^23^ evaluating the ID NOW platform (Abbott) had an original design to enroll 200 positive patients, but a prevalence drop made the study incompletable. Given the heterogeneity of the trials, finding a rigid, unifying criteria for inclusion was difficult and would defeat the purpose of this review.

In this work, as mentioned, our goal is reviewing the efficacy of tests in real life conditions. Therefore, we included studies that evaluated tests in patients in a true POC fashion and excluded laboratory-only or in silico evaluations. As a consequence, we have excluded from our analysis all proof-of-concepts and studies that used spiked samples. We have also excluded studies that used solely pre-tested frozen samples, as these conditions are vastly different from conditions in the field. As mentioned before, the selection of frozen samples may exclude samples with inhibitors, invalid or borderline results, and with low viral-loads, favoring samples with low Ct values.

Naturally, many trials used cooled or frozen samples at some point, especially when considering multi-platform evaluations. We tended to consider time from sample collection to testing in our selection criteria; while it was difficult to decide on a clear cut-off time, samples that were stored for a brief period of time to allow testing with a POC platform were accepted. We recognize this likely does not have the same value as fresh POC testing, it is often a necessary accommodation for validation studies where a comparator assay is used. As an example, Lephart et al collected samples from 88 patients (13 of which were known positive), stored at 4 °C and tested within 24h; this study was included in our table. On the other hand, studies using samples that were part of frozen panels tested in retrospect, often weeks after sample collection, were excluded. As an example of our criteria, we did not include the work by Corman et al^24^ who conducted a comparison of seven SARS-CoV-2 antigen tests available in Europe because processed samples were used and only negative swabs were collected from patients. We favored prospective studies.

Given the heterogeneity of designs and the conditions for studies, we debated between authors before inclusion when methods were not clear. For instance, Cerutti et al conducted a trial with 330 patients including a minority of frozen samples (n = 13); this trial was considered for our study^25^. Conversely, a prospective study by Courtellemont et al^26^ was not included as known positive patients were pre-selected to enroll; therefore, operators knew the status of the patients beforehand. A similar situation was found in the trial by Ghofrani et al^27^, who conducted a comparison with known positive patients and selected eligible samples; the methodology of this study was unclear and the sensitivity, reported as 96.7%, was much higher than usually reported in literature for antigen assays.

The vast majority of studies used nasopharyngeal samples, although a few studies used nasal samples only. Studies evaluating POC assays using saliva samples were reported^28, 29^ and usually show poor performance. For instance, Basso et al^30^ found a sensitivity of only 13% testing saliva with antigen tests; similarly, the performance of saliva samples was inferior in the study by Agulló et al^31^ evaluating the Panbio assay. We therefore reported the results for either nasopharyngeal or nasal swabs and (as per manufacturer’s advice) when they were part of an assessment with multiple sample types. When collection methods were compared (e.g, sensitivity of self-swab against healthcare collection^32, 33^), we reported results obtained by the healthcare staff worker.

We also excluded studies where POC assays were compared to other POC assays, as we considered no gold-standard was included. An example is the study conducted by Basu et al^34^, where Abbott ID Now COVID-19 was compared to the Cepheid Xpert Xpress SARS-CoV-2 without a PCR standard; the same can be said of the study conducted by Agarwal et al, which compared the Standard Q COVID-19 antigen test to the TrueNat POC platform^35^. It is important to mention that the reliability of the gold-standard was questioned in some studies^36^. Ideally, a reference standard would be built using a combination of criteria, including more than one assay^15^, cross-checked clinical information, radiological evidence and other laboratory information (e.g antibody production, viral markers or inflammatory makers), but this is understandably complex and unfeasible in many circumstances.

We also carefully considered the population type in the studies. Naturally, testing known-positive patients presents a bias. Due to challenges imposed by factors such as lockdowns, the urgency for results, different prevalence levels, differences in viral load at different stages of disease and the size of the trials, some studies tested known positive populations in order to obtain statistically significant data for sensitivity. If a study was conducted solely with known positive patients, it was excluded. On the other hand, studies that complemented a prospective evaluation by randomly testing positive populations were accepted. As an example, Basso et al tested antigen assays in 139 selected inpatients (this population had a 60% positivity rate) and 96 outpatients prospectively (3% positivity rate); this study was included. Some studies tested exclusively in a paediatric population, and were also included^37^.

Taking these factors into consideration, trials were assessed on the overall level of heterogeneity in their methodology. In the study conducted by Osterman et al^38^ evaluating 2 antigen assays, there was a significant variability between 2 sites as samples were collected in different time-frames (site 1 from March to October 2020 and site 2 between November-December 2020) and some samples in site 1 had different storage methods, with some being frozen for days before testing. Ultimately, we decided to include this study. Other studies like Marti et al^39^ were excluded due to a high level of heterogeneity in their methods, using both a POC and a centralised PCR as their standard and using different populations, including a population of known positive individuals. We attempted to include detailed explanations of the reasons for inclusion or exclusion of individual trials in appendix 1 of this review. We encourage authors to contact us for clarifications and potential corrections in the next version of this living review.

#### 2.33 Technical information on platforms

We investigated package inserts, instructions for use, comments on published studies and manufacturer’s websites in order to assess feasibility of POC placement and additional information that may be relevant to the end user. Apart from sensitivity and specificity, we included time to results, hands-on time, kit storage, machine operating conditions and regulatory status. We also made comments on testing requirements and additional details that were deemed relevant. We opted to use publicly available information, such as the instructions for use in the FDA website^13^. When that information was unavailable, we attempted to obtain the package insert by contacting the manufacturers or examining public information published by hospitals, government entities and other third-party institutions using the platform.

## 2. Results

**Figure 1.**
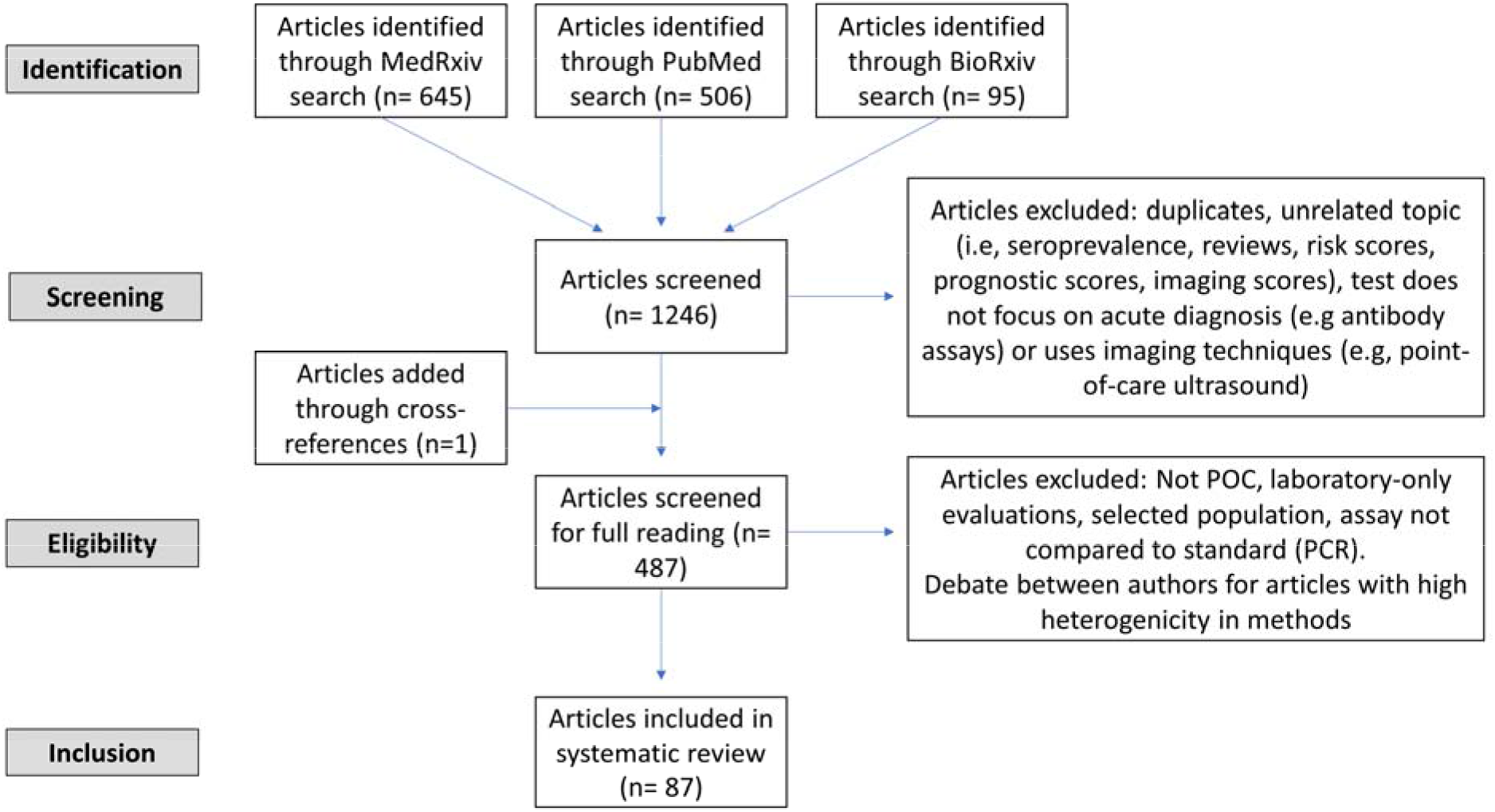
identification, screening, eligibility and inclusion flowsheet

### 3.1 Excluded studies and studies debated between authors

Comments on individual studies and their criteria for exclusion can be found in Appendix 1.

### 3.2 Comments on reference standards, sensitivity, specificity, positive predictive value (PPV) and negative predictive value (NPV)

POC tests have to be compared against a selected gold-standard in order to properly evaluate performance. This standard is real-time PCR in the vast majority of the studies (as mentioned above, some studies used another POC assay as a comparison and were excluded from our analysis). It is important to understand that the performance numbers reflect values against a reference standard, which is not always necessarily better or more accurate than the object of study. Most studies used a third platform as a tie-breaker in this context and the results of this full analysis were considered whenever appropriate. As mentioned previously, if a clear reference standard was used based antibodies and/or clinical and radiological evidence, this was also taken into consideration.

Positive predictive value (PPV) and negative predictive value (NPV) are useful to understand how much a result can be trusted given the prevalence of an infectious disease in a given time. Assays are always trialed settings with an estimated prevalence at a moment in time; in performance studies, the reference method provides that value. PPV and NPV change accordingly depending on the prevalence in the setting. For instance, the study of an antigen test by Peña et al^40^ reported a sensitivity of 69.86%, a specificity of 99.61%, but a PPV of 94.44% and a NPV of 97.22% as the prevalence in that setting was 8.64%. Here, we avoided using PPV and NPV projections whenever possible and aimed to report the provided ‘sensitivity’ and ‘specificity’ values, even though we recognize these values are intertwined. The decision to not use PPV and NPV projections was made because (1) in rapidly contagious infections like SARS-CoV-2, an accurate real-time monitoring of prevalence parameters is difficult, in contrast to what is found for infectious agents with a clearly defined and predictable epidemiology; therefore the prevalence of SARS-CoV-2 infection in a given setting may change rapidly considering outbreaks, lockdowns, and new variants. (2) An accurate real time monitoring of SARS-CoV-2 regional prevalence is challenging even for developed countries, resulting in prevalence values that are often retrospective. (3) An assay’s performance can be distorted using different prevalence levels. Finally, (4) an analysis including multiple PPV and NPV projections would make this review more speculative and less practical. We strongly suggest that the referenced trials are read in full for further information and clarification of performance as the term ‘sensitivity’ and ‘specificity’, as reported in this review, are always relative to prevalence in the particular setting of each study.

### 3.3 Included studies

We present the included studies in tables below. NAAT are grouped separately from antigen tests and are presented with an additional column discussing testing requirements. While most manufacturers used viral RNA copies/ml in a serial dilution to assess the assay limit of detection, some manufacturers used plaque forming units (PFU) instead of viral RNA copies/ml. We presented the limit of detection in copies/ml if both information were available, according to manufacturer’s instructions for use. The limit of detection of the different assays was converted into a copies/ml format when possible (for instance, if this value was given in copies/uL). We also did not include a claimed limit of detection for antigen assays.

We have not presented results for different populations (eg, symptomatic vs asymptomatic, children vs adults); therefore, the values presented represent the average for all individuals tested as provided in the studies. For example, the study by Ford et al^41^ presented separate values for children and adults, but we have reported these values as an overall.

Some assays had little technical information available, despite our best efforts to obtain package inserts or instructions for use. This was especially true for novel antigen tests. We have therefore included information to the best of our knowledge and indicated the information we could not obtain as “not available (N/A)” in the table.

Tables are divided between assays that diagnose of SARS-CoV-2 alone and multiplex assays (e.g, SARS-CoV-2 + FluA/FluB).

#### 3.31 SARS-CoV-2 studies

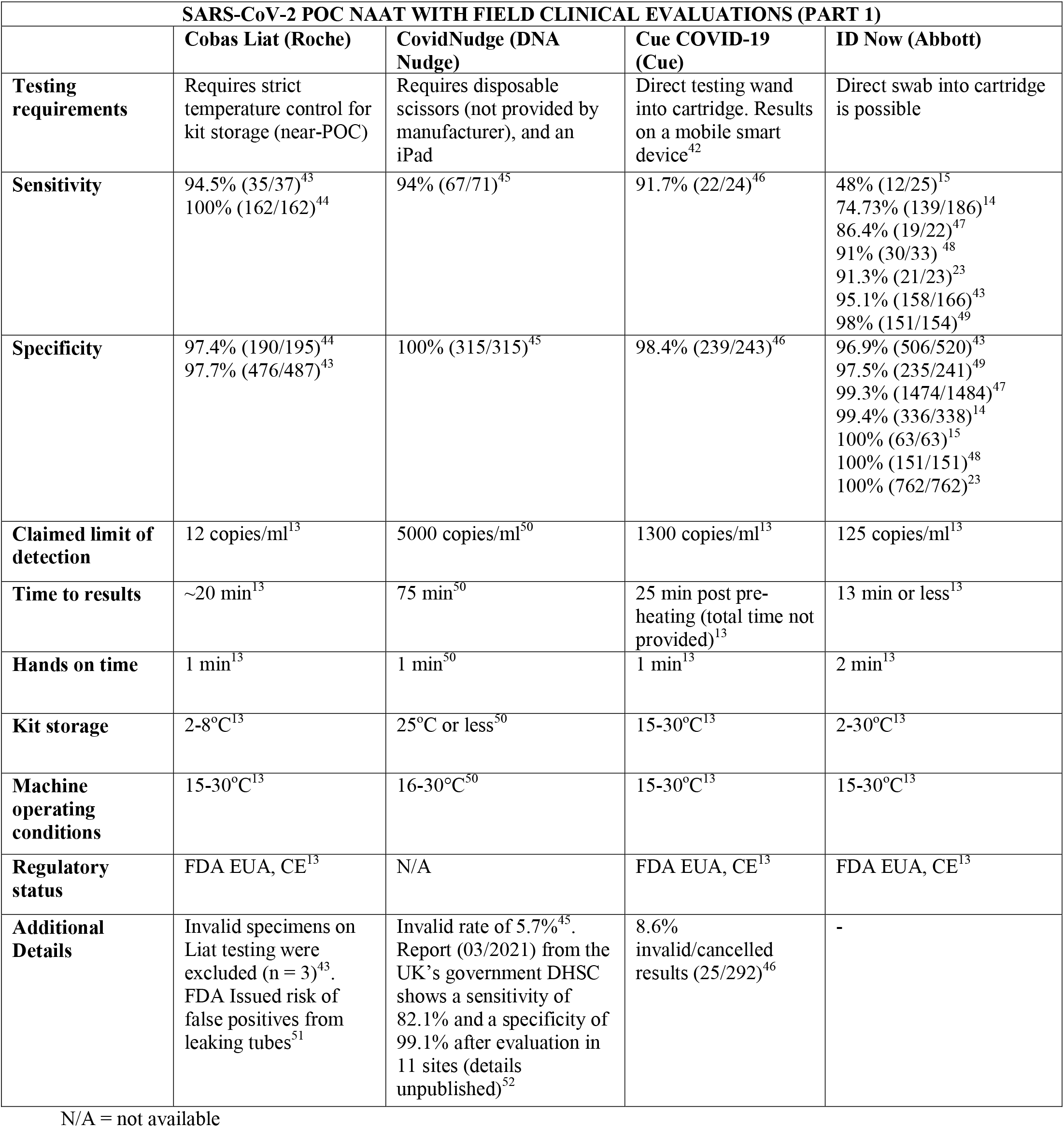
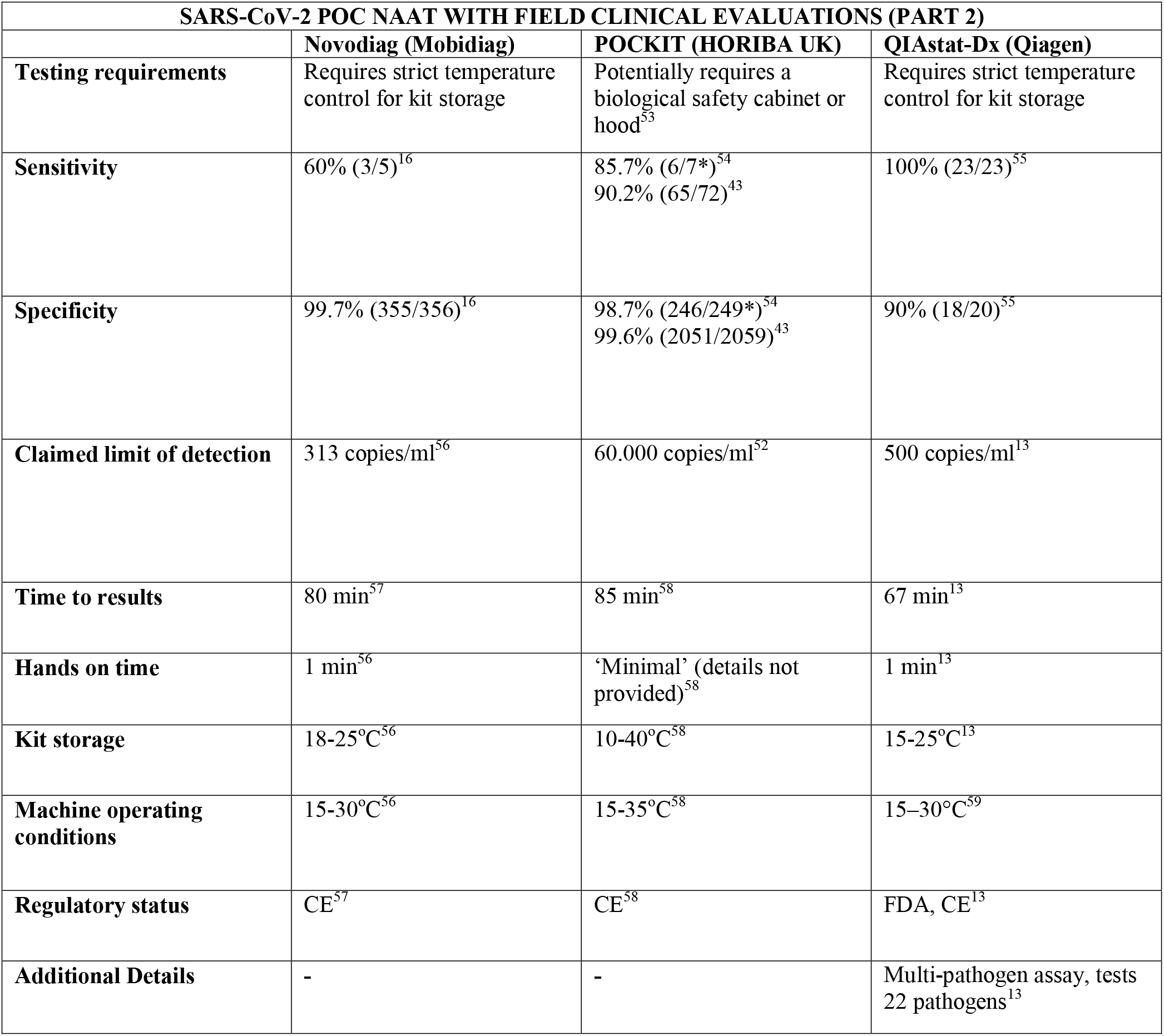
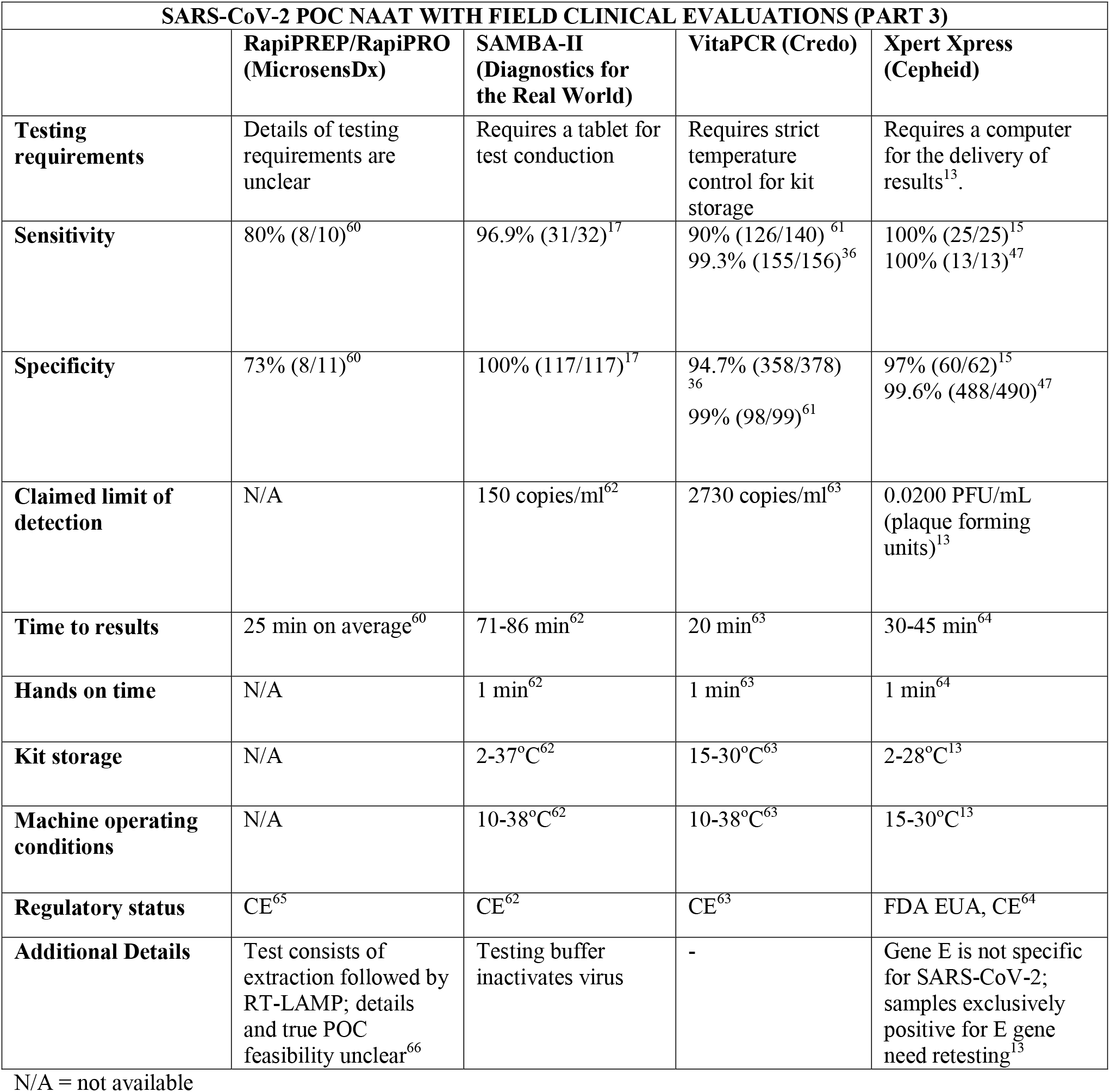
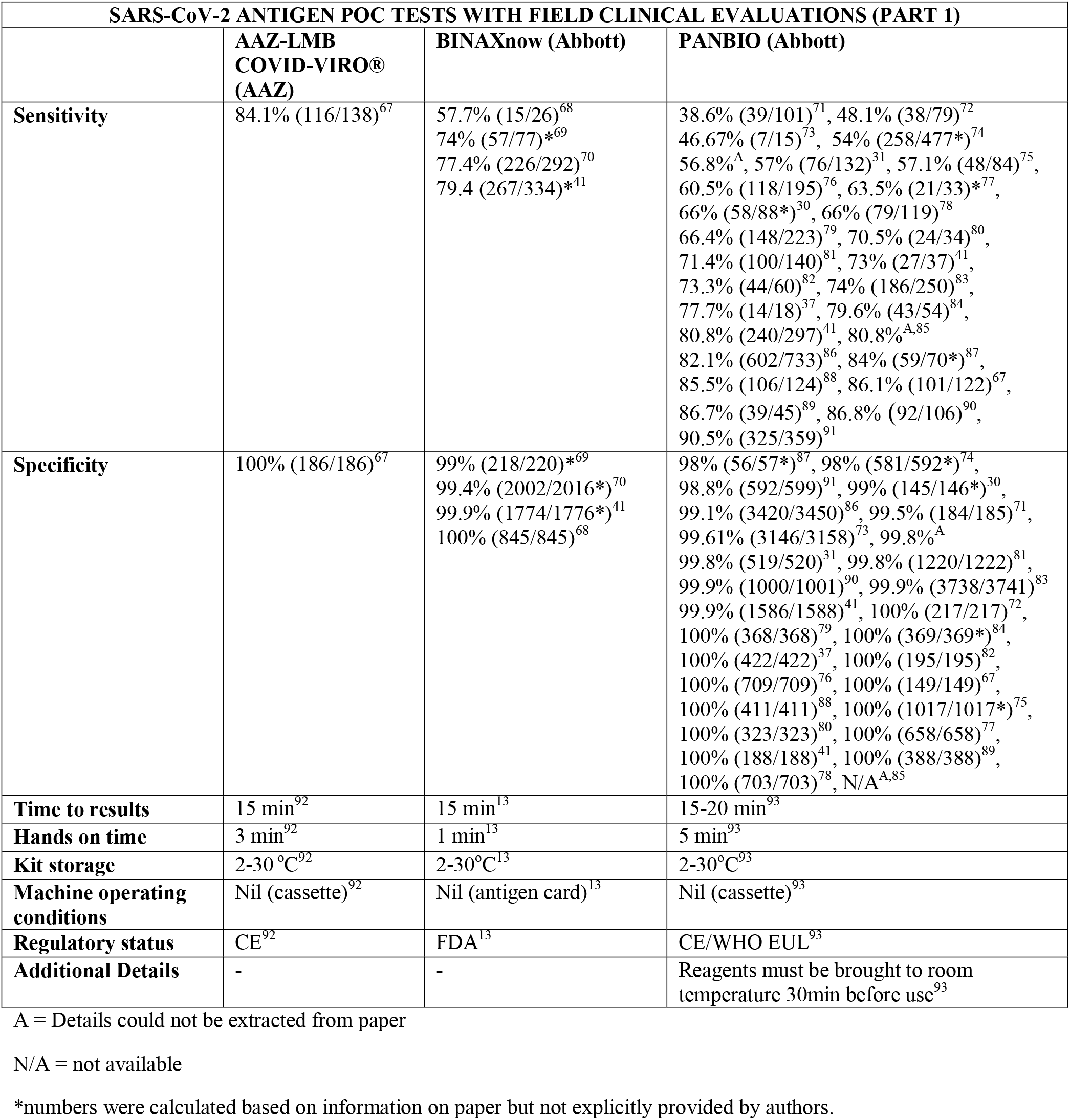
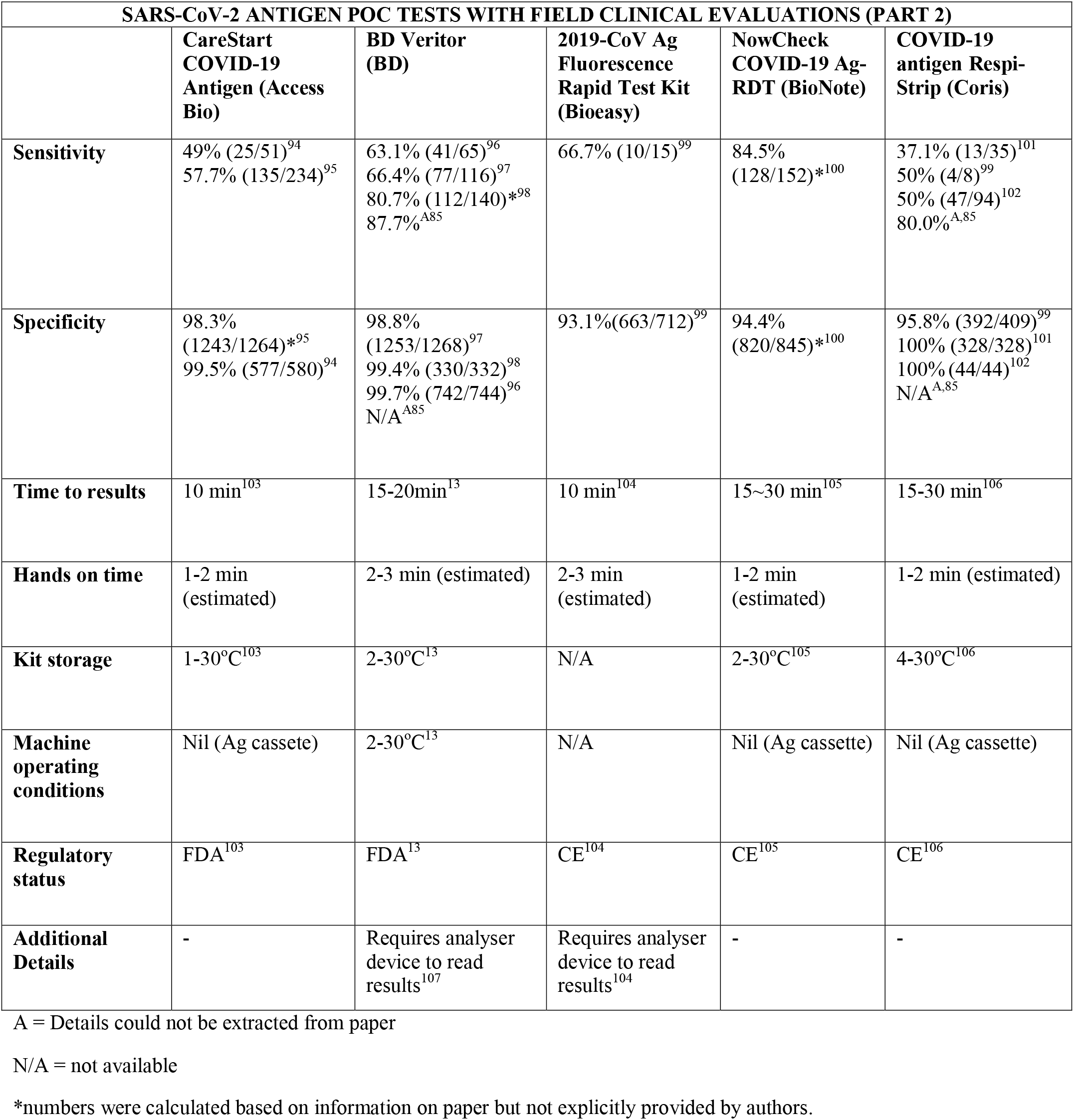
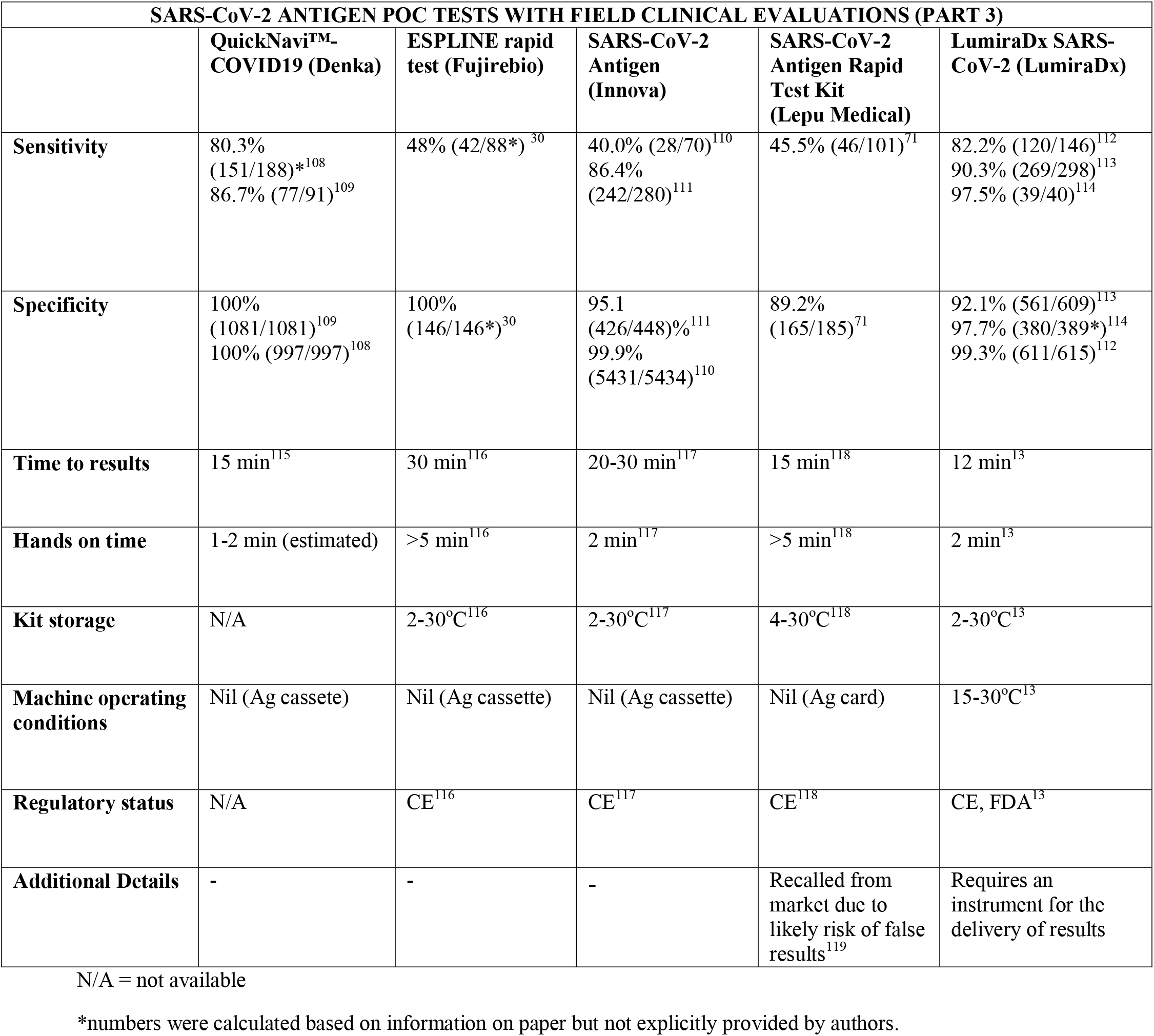
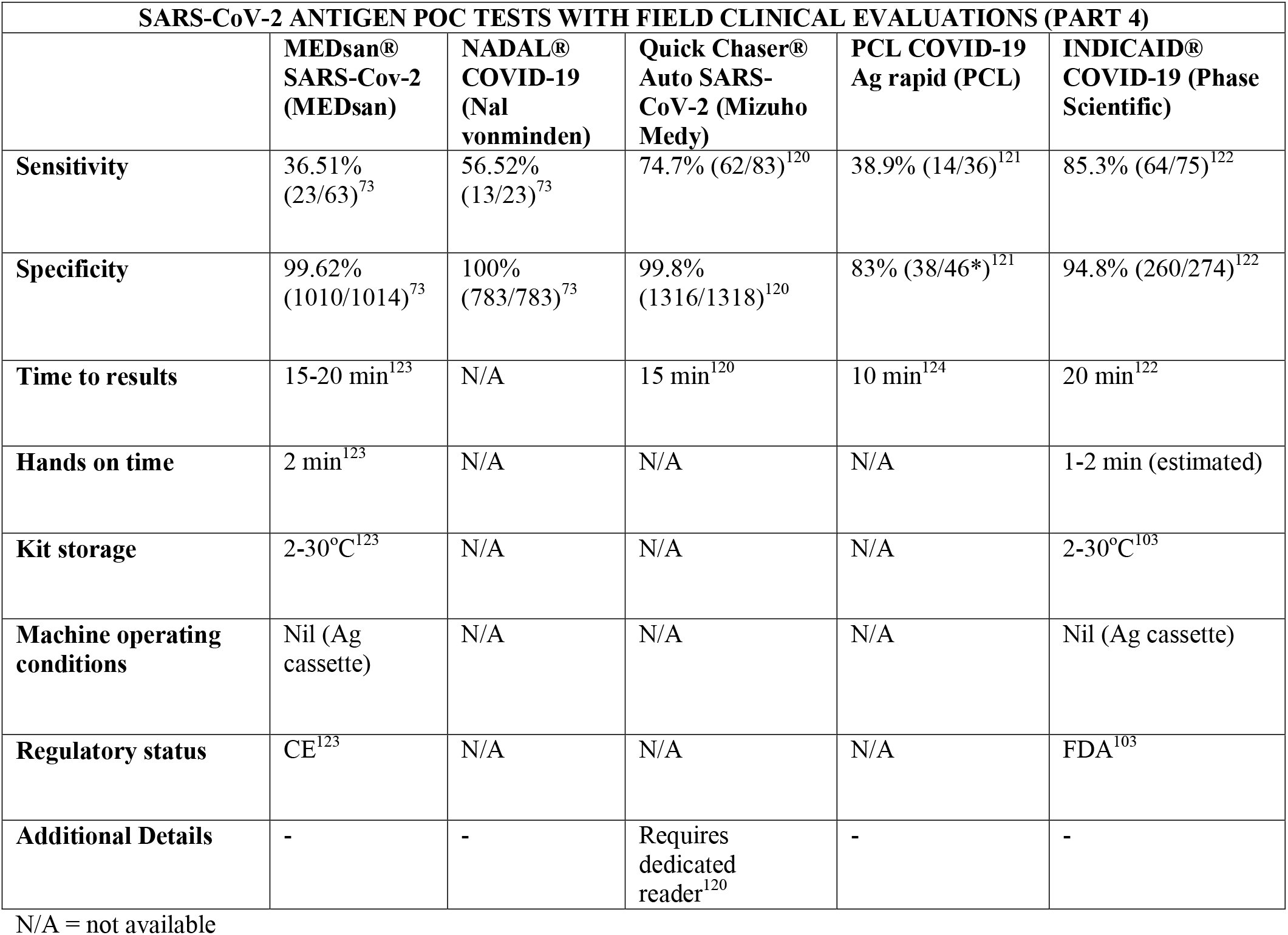
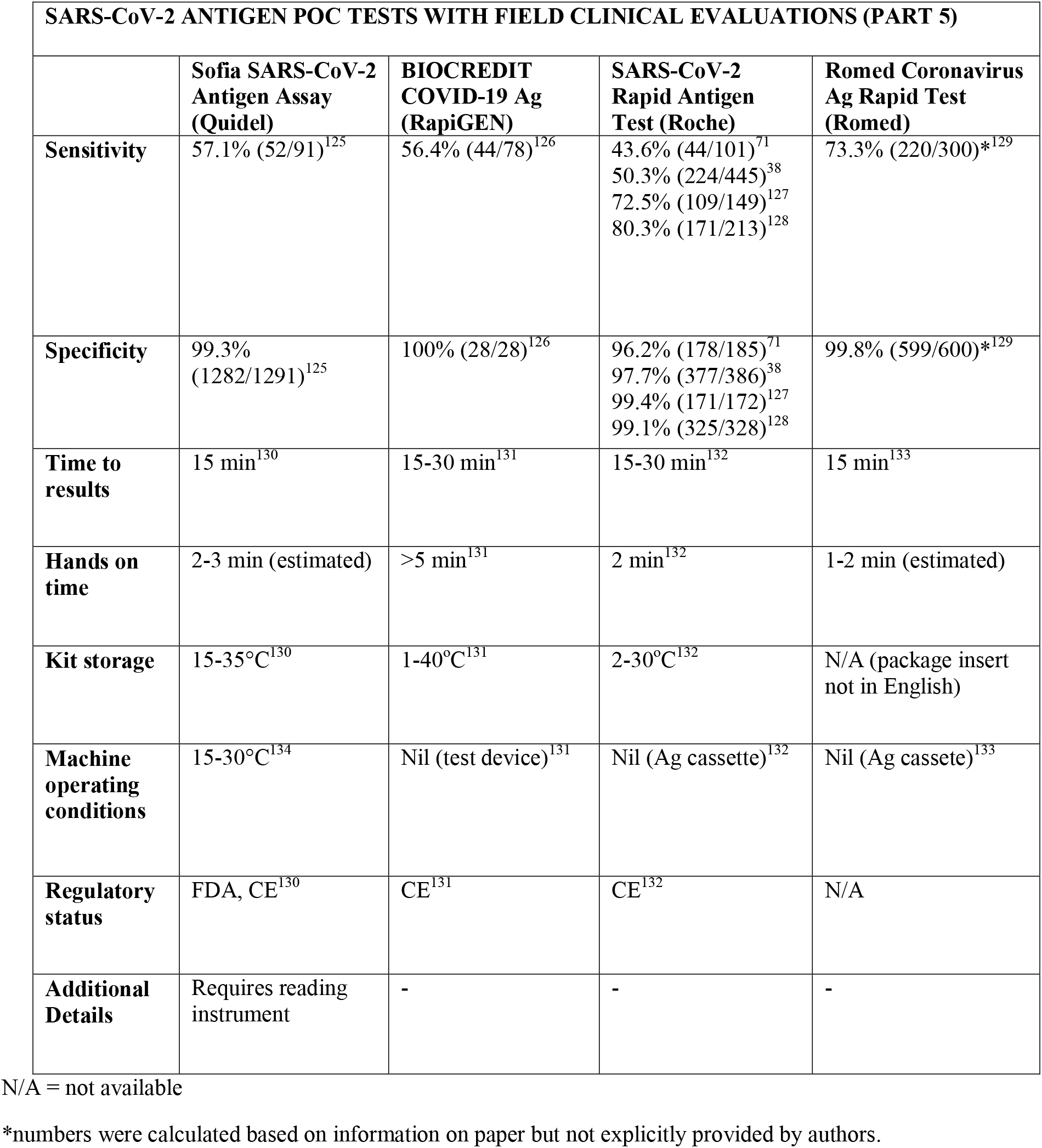
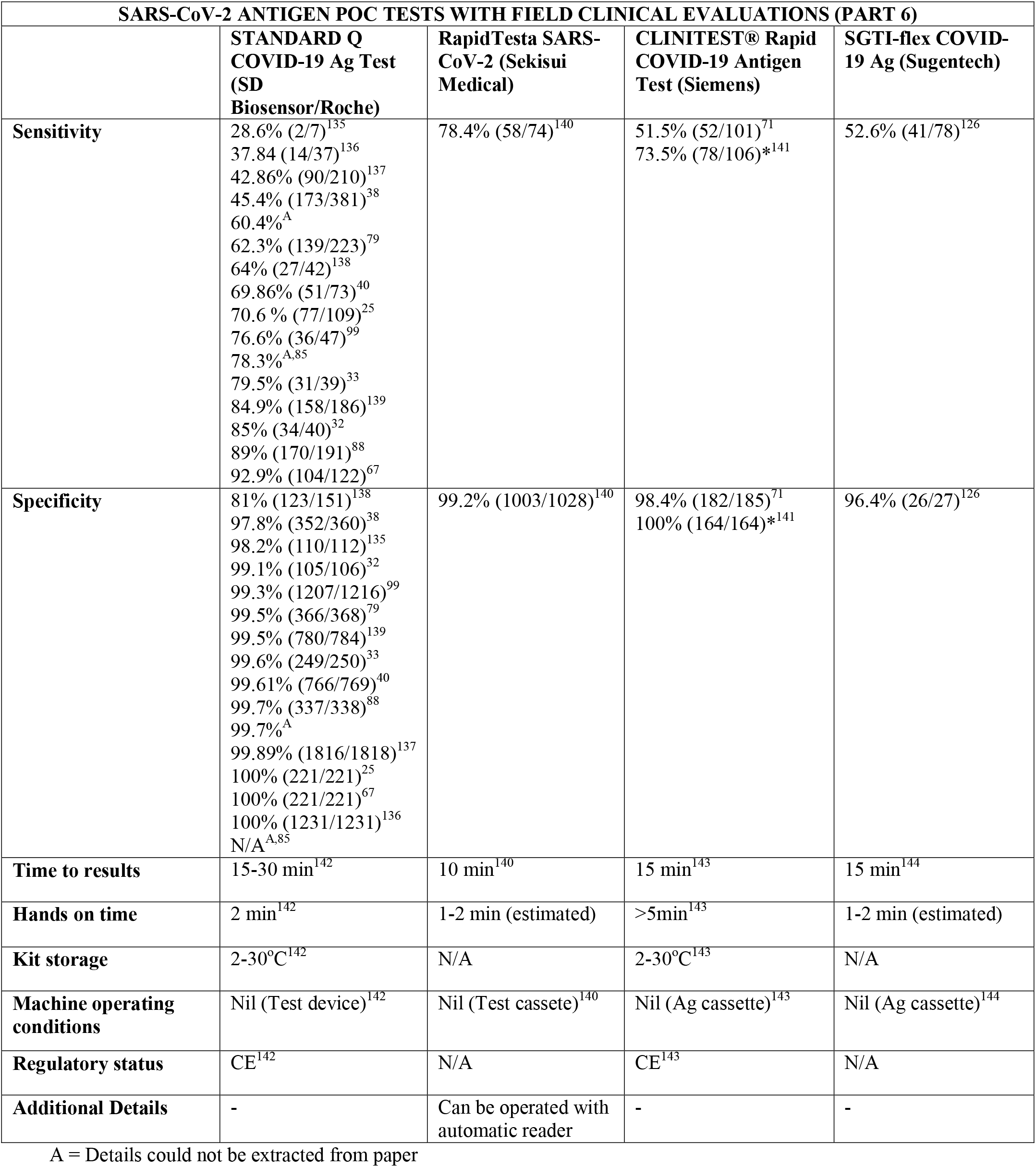

#### 3.31 Multiplex studies

Further adjustments to inclusion criteria had to be made to consider multiplex assays studies, taking into consideration factors like the drop in prevalence of some pathogens during the coronavirus pandemic, the different prevalence of pathogens across the globe and, consequently, the difficulty in getting enough numbers for a prospective analysis of pathogens other than SARS-CoV-2.

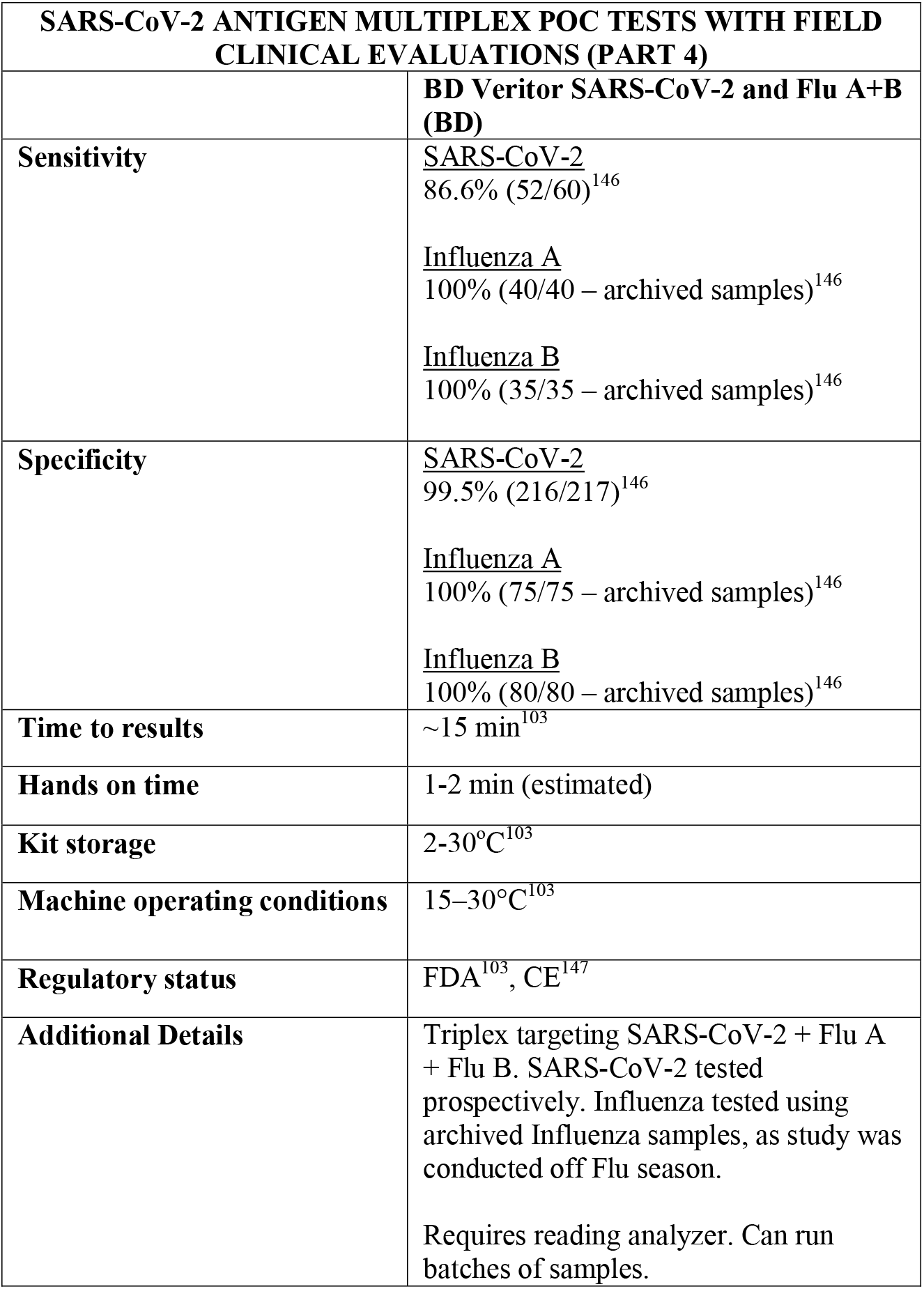

| **SARS-CoV-2 ANTIGEN MULTIPLEX POC TESTS WITH FIELD CLINICAL EVALUATIONS (PART 4)** |  |
| --- | --- |
|  | **BD Veritor SARS-CoV-2 and Flu A+B (BD)** |
| **Sensitivity** | <u>SARS-CoV-2</u><br>86.6% (52/60)<sup>146</sup><br><u>Influenza A</u><br>100% (40/40 – archived samples)<sup>146</sup><br><u>Influenza B</u><br>100% (35/35 – archived samples)<sup>146</sup> |
| **Specificity** | <u>SARS-CoV-2</u><br>99.5% (216/217)<sup>146</sup><br><u>Influenza A</u><br>100% (75/75 – archived samples)<sup>146</sup><br><u>Influenza B</u><br>100% (80/80 – archived samples)<sup>146</sup> |
| **Time to results** | ~15 min<sup>103</sup> |
| **Hands on time** | 1-2 min (estimated) |
| **Kit storage** | 2-30°C<sup>103</sup> |
| **Machine operating conditions** | 15–30°C<sup>103</sup> |
| **Regulatory status** | FDA<sup>103</sup>, CE<sup>147</sup> |
| **Additional Details** | Triplex targeting SARS-CoV-2 + Flu A + Flu B. SARS-CoV-2 tested prospectively. Influenza tested using archived Influenza samples, as study was conducted off Flu season.<br>Requires reading analyzer. Can run batches of samples. |

## 3 Discussion

It is well known that results from laboratory-only evaluations often differ from results of external clinical studies. Usually, evaluating an intervention in patients in real-life scenarios has an increased level of complexity. In the particular situation of SARS-CoV-2 testing, there are many possible reasons for those discrepancies. The first group of differences stems from factors related to the patients themselves, such as tolerance to swabbing and the presence of inhibitors in the nasopharynx secretions. There has been debate regarding the reliability of viral load and Ct values^148^, suggesting that viral load may greatly vary not only with the stage of the disease but also depending on the quality of swab collection and elements like dilution of swabs in a buffer and the RNA extraction process. Therefore, it appears to be important for a test to have a low limit of detection regardless of the ‘average’ viral load in a group of patients in order have a reliable diagnostic performance. The presence of inhibitors on samples is also relevant, as many assays describe interactions between food, beverages and medications and their amplification chemistry in their instructions for use or package inserts^13^. Once a sample is already known positive (with a known Ct value or an estimated viral load) and has been selected in a panel for a comparison against other test, this risk of poor-swabbing technique and inhibitors become non-existent, and therefore laboratory only evaluations may report higher sensitivity than what would be expected in a clinical setting.

Another group of differences is related to the feasibility of the workflow of the proposed testing platform in a true POC context, including the technique of swab collection and the expertise needed to conduct testing (such as sample preparation, instrument operation, and cleaning). Many tests that claim the possibility of use in a true POC setting would find resistance to adoption due to technical complications, as it is the case of most Loop mediated isothermal amplification (LAMP) assays (e.g, Yamazaki et al^149^). There are also factors related to the particularities of the pandemic scenario and the precautions needed to prevent cross-infections, including the desirable use of viral inactivation methods in samples and the suitability of amplification techniques that have the potential to release amplicons in a clinical setting, thus possibly increasing false-positive results. Furthermore, there are differences between laboratory evaluations and clinical settings; pre-selection of samples and repeating experiments is possible in a laboratory setting, but much harder inside a clinical workflow. In addition, the risk of false-positives in hospital environments is elevated, especially in areas of high movement and turnover like emergency departments, outpatient settings and clinical wards.

We identified 27 antigen tests and 11 NAAT with clinical trials that meet our defined criteria. These 38 assays were covered by 87 studies, with some studies covering more than one test. Considering the high heterogeneity of methods and outcomes between studies, and also the unbalanced number of studies per test, we opted not to conduct a meta-analysis in this study. We decided against providing an ‘average’ performance for tests as this would likely be misleading and would potentially downplay methodological discrepancies between trials.

NAAT, on average, take longer to provide results and require more equipment for test operation compared to antigen tests. However, NAAT results appear to be more reliable in clinical practice. Applying selection criteria specifically targeted at prospective studies, we noticed important differences between performance reported by manufacturers, laboratory evaluations and performance in field conditions. While this is true for both NAAT and antigen assays, the discrepancies were more marked in the antigen group. Healthcare facilities, individuals and test providers must be aware of the field performance of the tests before deciding on their implementation. We hope this systematic review can help making informed decisions regarding SARS-CoV-2 testing.

The accuracy of diagnostic tests is affected by numerous factors, including days since symptom onset, individual viral load, quality of sample collection, site of sample collection (nasopharyngeal, nasal only, pharyngeal only, saliva only) and test modality (nucleic acid amplification or antigen). As previously mentioned, all studies included were compared to PCR or qPCR assays, considered the gold standard for SARS-CoV-2 diagnosis. Results suggest a strong tendency of antigen tests to be less accurate than NAAT in field clinical trials. This finding is aligned with findings of other reviews^20^.

Importantly, we noticed differences in performance within a given test. For instance, the sensitivity of the antigen assay STANDARD Q COVID-19 Ag Test (SD Biosensor) ranged from 28% to 92%, and the sensitivity of the antigen test PANBIO (Abbott) ranged from 38 to 90%. The explanations for the phenomenon of high variability in results, particularly in sensitivity, are likely multifactorial, including differences in SARS-CoV-2 prevalence between studies and differences in sample size and methodology. One such factor may be testing kits transporting and storage, as most antigen tests need to be stored between 5 and 30C°. Haage et al^150^ assessed 11 antigen tests and identified reductions of up to 10 fold in sensitivity for 46% of the assays after 10 minutes outside the ideal temperature range; this number grew to 73% if the exposure lasted three weeks. Pollock et al^70^ reported similar findings evaluating the BinaxNOW test. This finding is significant and may partially explain false negative results, considering that many regions have oscillations in temperature outside the target range and some factors like stock storage and transportation are beyond the end-user control. In this context, Abdul-Mumin et al^138^ reported suboptimal results for both sensitivity and specificity using the STANDARD Q COVID-19 test (64% and 81%, respectively); authors reported the difficulty of maintaining the tests within the ideal temperature range near patients given the temperatures in Ghana, despite following the manufacturer’s storage conditions. Therefore, for optimal results, it is important to consider not only the performance of the tests but also context they are being used in.

Point-mutations generating changes in the SARS-CoV-2 nucleocapsid protein structures can also play a role in suboptimal performance; Bourassa et al uncovered a 1000-fold loss in sensitivity for the Sofia Antigen test (Quidel) which was associated with the D399N mutation^151^. Importantly, the high variability range in results may reflect a publishing bias, as high variability was usually seen in assays with the largest number of published studies.

Additionally, we found some evidence that the sensitivity of antigen tests increases if they are used within the first days of symptoms, but this is still significantly inferior to the average performance of NAAT. For instance, Bulilete at al reported that the sensitivity of the Panbio assay improved from 71.4% to 77.2% if the test was conducted in the first 5 days of symptoms^81^. However, using time from symptoms to choose between test modality has several limitations. The definition of being symptomatic and date of first symptom can be subjective, depending on factors such as threshold of perception and memory, which not only may vary between individuals but may also be unreliable. This is especially important in settings such as care-homes, accident and emergency rooms and for populations such as children and cognitively impaired individuals. Furthermore, respiratory compromise is usually a late clinical presentation of COVID and therefore it is reasonable to expect a significant portion of patients to present late to healthcare services, when symptoms worsen. Not only memory of symptom onset may be weakened by then, but a significant portion of frail individuals may also present with delirium. At that stage, timely interventions such as the use of dexamethasone in patients requiring respiratory support will require confirmation of SARS-CoV-2 presence^18^, which can be challenging if tests only perform well for a small period of time. In addition, there is evidence that immunosuppressed individuals, who are more likely to have complications of SARS-CoV-2 infection, may have different viral load and shedding patterns, making the use of time criteria for diagnosis even more complicated^152^.

Some studies have also provided different performance values for symptomatic and asymptomatic individuals, but a division between those two groups of individuals has similar problems with symptom recognition. Also, a significant portion of asymptomatic individuals are in fact pre-symptomatic and will develop symptoms in the future, but may already be in the shedding phase; this becomes even more important if the individual has had a high-risk contact.

It appears that around the 8^th^ day after the beginning of symptoms, individuals are not capable of transmitting SARS-CoV-2^153^. It has also been proposed that antigen tests or Ct values of PCR tests could be used as a cut-off for infectivity. This approach assumes that individuals with a negative antigen test or a high Ct value would hardly have a viral load capable of kickstart infection in another individual. While information such as Ct values can be informative, there may be several caveats to the use of Ct values for clinical decision making. Factors such as the quality of swabbing and the tolerance of the patient can add intrinsic variability to estimations of viral load (and thus Ct values), as well as issues with transportation of samples and limitations of the extraction and amplification systems^148^. As Binnicker discusses, even assays using the same gene target may have significant variations in Ct value^154^, and factors such as the type of sample and swab also need to be considered. Additionally, the volume of buffer the swab is diluted into and the transport media itself can also affect viral load, and it is likely that the quantities of virus in the nasopharynx surfaces are uneven, varying between different regions of the tract and depending on factors such as the ingestion of food and drinks and time. As Basso et al has demonstrated, repeat testing of the same sample can result in different Ct values^155^, and this variability has been reported in clinical practice as well^156^. It is also unclear which Ct value would be used as a cut-off for infectiousness, as values as different as 24^157^ and 34^158^ have been proposed in the literature.

Some studies also showcased the implications of using tests with suboptimal specificity in settings of low prevalence. Hoehl et al^159^ used an antigen test for self-collected home testing of teachers, with the goal to prevent clusters of infections. Out of a population of 602 individuals, 5 were confirmed positive but 16 false positive results were recorded. The same concern was voiced by Kriemler et al^160^ when using antigen tests to assess the point-prevalence of acute SARS-CoV-2 infections in school children. In a study by Colavita et al, of 73,634 individuals tested at international airports, 1176 were reported antigen positive but only 34.3% were confirmed positive. A post-implementation assessment of two antigen tests used for screening of asymptomatic staff (n=71,847) conducted by Kanji et al^161^ had similar findings.

Regarding kit storage, most platforms will require the use of refrigerated or air-conditioned facilities given the average upper storage limit was 30°C. Some of them deserve mention for requiring strict temperature control, particularly the Cobas Liat (2-8°C), the QIAstat-DX (15-25°C) and Novodiag (18- 25°C). Only 5 assays have a published kit storage temperature above 30°C: SAMBA-II (NAAT), VitaPCR (NAAT), HORIBA (NAAT), Sofia SARS-CoV-2 (antigen) and BIOCREDIT (antigen).

Time to results was highly variable between NAAT, ranging from ∼13 minutes (ID Now) to ∼85 minutes (HORIBA, SAMBA-II). Antigen tests take an average of 15 minutes for results, not surpassing 30 minutes. Hands-on time, used in this context as the time needed to prepare the test before a run (prepare samples, load machine, configure test conduction) was usually around 1 to 2 minutes, and rarely over 5 minutes across all platforms.

### 4.2 Other assays

Assays other than NAAT and antigen tests have also been used for COVID-19 diagnosis or SARS-CoV-2 identification. We found a few studies using the FebriDx device (Lumos diagnostics), which captures Myxovirus resistance protein A (MxA - a marker of interferon-induced antiviral host response) and C reactive protein (a well-known and widely used inflammatory marker in medical practice). In one study, the assay had a sensitivity of 93% and a specificity of 86%^162^ (with an estimated prevalence of 48% in the studied population). There are other studies available regarding this assay^163–165^ but a comprehensive analysis of this platform goes beyond the purpose of our review. As the markers are commonly elevated for a range of pathogens, the test has a low specificity and has limited use in settings with low prevalence.

Few tests had a satisfactory number of clinical studies, and in many situations the number of individuals enrolled was suboptimal. Further research and reviews of this topic are encouraged.

### 4.3 Limitations

One of the main limitations of this review is the selection criteria. Considering the high heterogeneity of methods and outcomes between studies, finding a clear-cut unified exclusion criteria was not possible. We debated between authors when in doubt, but a level of subjectivity was inevitable.

For the same reasons of heterogeneity, we opted not to conduct a meta-analysis in this study. Authors have decided against providing an ‘average’ performance for platforms as this would likely be misleading and would potentially downplay the method discrepancies in the trials.

## Data Availability

No special characters

## 5. Other information

### 5.1 Registration

This review was registered in the International prospective register of systematic reviews (PROSPERO) with registration number CRD42021260694. A protocol for this study can be assessed online.

## Appendix 1 comments on individual studies and exclusions

In this section, we comment on criteria for inclusion or exclusion of individual studies. As the criteria for exclusion can often be similar, not all excluded studies will be mentioned. We aim to provide commentary especially when inclusion or exclusion was debated between authors and to give examples of our exclusion pathway.

Multiple studies were excluded in our final screening as tests were conducted using spiked or synthetic samples. The study by Zheng et al^166^ evaluating an unnamed lateral flow dipstick assay is an example. Another example is the work of Tanida et al^167^, evaluating the ARIES SARS-CoV-2 Assay; this study used patient samples with defined copy numbers and synthetically spiked samples. The study by Shelite et al^168^ was not included as it was conducted with processed, archived samples. Details of the platform and it’s true feasibility for POC placement are unclear.

As the case of most LAMP assays, the saliva LAMP assay proposed by Yamazaki et al^149^ was not deemed to be feasible in a true POC scenario as it requires RNA extraction, a heat block and apparently multiple pipetting steps, and therefore a reasonable level of expertise. The assay was also tested with pre-selected samples. The study of Desai et al^169^ evaluating the Atila iAMP test and the OptiGene Direct Plus RT-LAMP was not included as platforms were also not considered feasible for POC implementation. The study by Yoshikawa et al^170^ and Casati et al^171^ are also examples of this exclusion criteria. The study by Nörz et al^172^ evaluating the Elecsys® SARS-CoV-2 Antigen assay was not deemed feasible in a POC environment given the modules necessary for test conduction are centralised equipment with a large footprint.

Similarly, the study conducted by Peto et al^173^ regarding the LamPORE platform was not included in the analysis as the platform was not deemed to be feasible for POC implementation. RNA needs to be extracted and primers have to be added and incubated with a thermocycler, followed by multiple manual steps. The trial also was conducted with pre-selected, frozen samples. For similar reasons, the study by Singha et al^174^ using a glucose meter to detect SARS-CoV-2 was not included, as the test requires a centrifuge, a magnet, and incubation in water baths. This evaluation was also conducted with known positive samples only.

The study by Caffry et al^175^ using the Q-POC device was not included as the study was conducted with known-positive/negative frozen samples. Another example is the study by Krause et al^176^ regarding the ID now platform, conducted with frozen, pre-processed samples. The study by Hagbom et al^177^ was also not included given samples were collected from known-positive patients. Similarly, the study by Muthamia et al^178^ evaluating the BD Veritor platform was not included as the analysis was conducted with known-positive patients. Nordgren et al^179^ was excluded given test was conducted with pre-selected, known positive samples. Onyilagha et al^180^ evaluation of the Biomeme SARS-CoV-2 assay and the Precision Biomonitoring TripleLock SARS-CoV-2 assay was not included given evaluations were made with known positive samples. A platform was also used for extraction, though this was considered to be feasible for POC use. Stokes et al^181^ evaluation of the PANBIO antigen test was not included as it only recruited known positive individuals.

The study by Renzoni et al^182^ evaluating the Visby Medical RT-PCR Portable Device was not included given it was conducted retrospectively, with frozen samples. Agulló et al evaluated nasopharyngeal, nasal only, and saliva samples against nasopharyngeal samples in the Cobas z 480 Analyzer (Roche). Because of this division, sample size ended up being small and heterogeneous. The concordance for positive results was 57.3% for nasopharyngeal samples and as low as 23.1% in saliva; we included the results of the nasopharyngeal testing in our table. Studies conducted with frozen samples are available for this assay^183^, but as mentioned, were not included in our review.

Basso et al^30^ tested antigen assays in both saliva and NPS in a mixed population (139 inpatients, 96 outpatients), providing individual figures of sensitivity and specificity for the NPS samples. Since the comparator gold-standard was also tested in saliva and NPS, the ultimate reference standard was unclear. In the case of antigen tests, the detailed number of individuals to give the figures for sensitivity and specificity were not provided in the study or in the supplementary material to the best of our knowledge. We decided to include this study in our table with a commentary pointing towards the fact that the number of individuals used to make the figures was an estimation made for practical purposes and was not provided in the original paper; therefore, it may reflect slightly different patient numbers.

Halfon et al^184^ study was not included as samples were pre-selected based on symptom onset and Ct value. Similarly, the study conducted by Jääskeläinen et al^185^ evaluating 3 antigen platforms was not included due to the use of pre-selected, known-positive frozen samples. The study conducted by Blairon et al^186^ was excluded due to the same reason. Kweon et al^187^ also conducted an evaluation of 2 antigen platforms where positive samples were obtained by testing known-positive individuals and pre-selected before the evaluation, and thus was not included in our results.

An unnamed antigen test by R-Biopharm was also evaluated by Toptan et al^188^ but this study was not included given that testing was conducted in archived samples only. In line the same criteria, the evaluation of the SIENNA™ COVID-19 Antigen test by Bouassa et al^189^ was not included as the comparison was made in the laboratory with frozen samples. The study by Mitchell and George^190^ evaluating the ID NOW assay and the study by Assennato et al^191^ evaluating the SAMBA-II platform were excluded for the same reasons. The study by Stevens et al^192^ evaluating the Cepheid Xpert Xpress SARS-CoV-2 assay was also not included given the analysis was made on frozen, pre-selected samples. Consequently, the study by Young et al^193^ assessing BD Veritor and the Sofia 2 SARS Antigen test was excluded as samples were shipped frozen and evaluations happened in a laboratory setting. Miscio et al^194^ evaluated the bKIT Virus Finder COVID-19” (Hyris Ltd); this study used frozen samples which were further manipulated, and therefore was not included in our list of trials. Another example is the study by Panpradist et al^195^ evaluating the Harmony COVID-19 platform. The study by Koskinen et al^196^ on the MariPOC platform (ArcDia) was not included as it was conducted retrospectively with frozen samples.

Some studies were excluded due to methodology. Hoehl et al^159^ study using the RIDA® QUICK SARS-CoV-2 (R-Biopharm) was not included as a minimal number of samples were tested with a confirmation method, and thus false negative results could not be determined. The study by Mlcochova et al^197^ was not included in our table because (1) the methodology used frozen pre-selected samples, (2) antibodies were evaluated together with NAAT, making the selected time-frame for analysis questionable as NAAT was used to test samples up to 28 days after symptoms, (3) the criteria used for the reference standard was not entirely clear and (4) the number of samples was small (n=45) and divided between different assays. For similar reasons, the study by Veyrenche^198^ et al was not included in our list. A study by Micocci et al^9^ evaluating the LumiraDx antigen test in care homes was not included due to methodology individuals were tested three times a week, twice a week with antigen tests but only once a week with a RT-PCR standard.

After discussion between authors, the study by Olearo et al^199^ that evaluated 4 antigen tests was not included in the table for having openly deviated from the manufacturers recommended sample matrix/handling instructions and presenting a sensitivity between 49.4-54.9%%, which is on the lower side of what is expected for antigen tests.

The methodology in the study by Hogan et al^200^ evaluating the Mesa Accula assay (now the Thermo Scientific™ Accula™ SARS-CoV-2 Test after acquisition by Thermo Fisher) was not totally clear, since there is no mention of frozen samples or time to test after sample collection. However, it appears that samples were pre-selected (N=100) and tested in a laboratory after being tested by a centralised PCR assay. We therefore believe that this study, which showed a sensitivity of 68% and a specificity of 100%, is unlikely to accurately reflect results in the field.

A different methodology was used by Rastawicki et al^121^ evaluating the PCL COVID-19 Ag rapid fluorescent immunoassay (FIA); 4 swabs were collected in the course of 2 days and antibodies were also evaluated. We considered the comparison between RT-PCR and antigen test straightforward enough for the trial to be included in our table, despite the low number of patients enrolled.

The study by Regev-Yochay^201^ could not be included as multiple antigen platforms were classified as a whole, and individual data for individual assays was not available.

The study by Smithgall et al^202^ regarding the Cepheid Xpert Xpress and Abbott ID Now was not included as only remnant patient samples were tested; therefore, the platforms were not evaluated in a proper clinical environment. Loeffelholz et al^203^ study evaluating the Xpert Xpress SARS-CoV-2 was excluded as all but one site tested the platform with remnant frozen samples. The study by Wolters et al^204^ on the same platform was also excluded as it only evaluated diluted and processed sample panels in a laboratory setting.

Jokela et al^16^ has made two different evaluations of the Novodiag by Mobidiag where, by our understanding, an initial phase was a laboratory evaluation and the second phase was a prospective clinical trial. However, there was a major drop in prevalence in the second phase of the study, which only enabled collection of 5 positive samples in a population of 362 individuals. We included the second phase of this trial in our table despite the low number of positive samples.

Moeren et al^98^ study evaluating the BD Veritor antigen test had a mixed design, where 352 symptomatic adults were evaluated prospectively and known-positive individuals (n = 123) were added to the pool, visiting them at home within 72h of their RT-PCR positive result. Because the assay was tested in a true POC fashion and this was necessary to obtain statistical significance, we decided to include this study in our table. We also reported the specificity considering the 2 false-positive results obtained by the analyser, not considering the eye readings.

The study by Berke et al on the Visby Medical COVID-19 test using pooling strategies in a school setting was not a performance study, as negative pools were not tested against a gold-standard. Therefore, it was not included in our study.

The study by Cassuto et al^205^ regarding the COVID□VIRO® antigen assay was not included given weak-positive samples were excluded from analysis.

The study by Kernéis et al^206^ was not included as we could not assess the true performance of the antigen test against NAAT. Authors report a positivity rate of 129/1443 for NAAT tests and 89/1115 for antigen tests, but further details are unclear. A reference standard that included saliva samples amplified with NAAT was used later, which further complicated our understanding.

The study by Lamb et al^207^ was not included as no sensitivity analysis was conducted; only positive individuals were offered a confirmatory PCR test. The study by LeGoff et al^208^ was not included; the goal was to evaluate a saliva LAMP test, and despite using an antigen test, not all RT-PCR samples had paired antigen samples.

Linder et al^209^ report on three antigen-detecting tests had no detailed methodology, and therefore was not included. The study by Perkins et al^210^ was not included given only 37 antigen tests were conducted for 853 individuals, thus making a performance analysis not adequate.

The study by Oshiro et al^211^ where an antigen test was developed had an unclear methodology, but followed a proof-of-concept model and to the best of our knowledge was not tested in a true POC fashion. Therefore, it was not included. The study by Papadakis et al^212^ was excluded for similar reasons; the evaluation without extracted RNA also used a high proportion of frozen samples compared to fresh samples. The study by Wu et al^213^ evaluating a newly developed cross-priming isothermal amplification kit (named Kit A) is another example.

The study by Wen et al^214^ evaluating the Xpert Xpress (Cepheid) platform was not included as it was conducted with pre-selected, known-positive frozen samples; the study mentions a small number of tests on fresh specimens (of which 6 were positive) but in addition to the proportionally small number, it is unclear if those specimens were integrated in the final analysis^214^.

Pilarowski et al^68^ has changed the criteria for evaluation of a positive result in the middle of the trial, later recalculating their data. This apparently diverged from manufacturer’s instructions for use but was advised by experts by the manufacturer’s research staff, and therefore we have included this study.

The study by Šterbenc et al^215^ evaluating the Rapid Antigen Test (Roche) was not included given its methodology was not optimised for an evaluation of performance. Only two positive samples were found in the small population (n = 36), tested three times a week for 2 weeks (191 samples); one of them was considered the tail capture of RNA (the individual was positive for IgG previously) and the other was missed by the antigen test, which would give a sensitivity of 0%.

The study conducted by Tulloch et al^216^ evaluating the Innova SARS-CoV-2 antigen test was not optimised to be a performance study; only 828 of the 1638 samples had matching PCR tests. We therefore excluded this study as the gold-standard was not employed systematically.

Osmanodja et al^145^ have trialed a test that was generically called ‘Novel SARS-CoV-2 Antigen-Detecting Rapid Diagnostic Test’, reporting a sensitivity of 88.6% and a specificity of 99.7%. Because we could not find further details on this platform, we have opted to not include it in the comparison tables.

